# A Sensory-Cognitive Dissociation in Listeners with Hearing Difficulties: An Exploratory Analysis Linking Tinnitus to Binaural Unmasking Deficits and Speech Complaints to Memory

**DOI:** 10.64898/2025.12.18.25342552

**Authors:** Stefan Bleeck, Yasmeen Hamza

**Affiliations:** Institute of Sound and Vibration Research, University of Southampton

## Abstract

**Background:** The construct of Hidden Hearing Loss (HHL) proposes a link between patient-reported hearing difficulties and underlying neural deficits not captured by the standard audiogram. However, the heterogeneity of this population challenges the utility of HHL as a unitary diagnosis. This study presents an exploratory analysis aimed at deconstructing the HHL symptom complex.

**Methods:** In 30 participants with a range of hearing abilities and complaints, we measured binaural unmasking using the Binaural Intelligibility Level Difference (BILD). We employed a two-stage analysis. First, a “lumping” analysis tested whether participants could be grouped into a unitary “HHL profile” that predicted a BILD deficit, using both theory-driven classification and data-driven clustering. Second, after this approach failed, a pre-planned exploratory “splitting” analysis used a Linear Mixed-Effects Model (LMM) to investigate whether individual clinical markers (tinnitus, self-reported speech difficulty) were independently associated with the BILD.

**Results:** The “lumping” analyses failed to find a significant difference in the BILD between subgroups, questioning the utility of a unitary HHL profile. In contrast, the exploratory “splitting” analysis found a significant interaction between tinnitus and listening condition (β = 1.57, p = 0.009), suggesting that participants with tinnitus exhibited a smaller BILD. The complaint of speech perception difficulty was not significantly associated with a BILD deficit (p = 0.086) but was associated with lower scores on a test of short-term memory (forward digit span, p = 0.046).

**Conclusion:** Our findings challenge the value of a unitary HHL profile for predicting this specific binaural deficit. Instead, our exploratory analysis generated a specific, testable hypothesis of a sensory-cognitive dissociation: in our sample, tinnitus was associated with a reduced capacity for binaural unmasking, while the complaint of speech difficulty was associated with poorer short-term memory. These preliminary findings, derived from post-hoc analysis of an underpowered study, require rigorous validation in larger, pre-registered studies.

## Introduction

Listeners who report difficulty understanding speech in noise despite having normal or near-normal audiograms present a significant clinical and scientific puzzle (Schaette & McAlpine, 2011). This condition, often termed Hidden Hearing Loss (HHL), is hypothesized to stem from neural deficits, such as cochlear synaptopathy or demyelination, that degrade the temporal precision of auditory signals without affecting absolute thresholds (Liberman & Kujawa, 2017; Moser & Starr, 2016). However, the HHL label groups together a wide range of individuals with diverse complaints, most notably tinnitus and general speech-in-noise (SiN) difficulty. The assumption that these disparate symptoms arise from a single underlying pathology and manifest as a uniform functional deficit remains largely untested.

This study was designed as an exploratory, hypothesis-generating investigation to challenge the coherence of the HHL construct. We acknowledge from the outset that our small sample size (N=30) renders the study underpowered for definitive, confirmatory conclusions. Instead, our goal is to deconstruct the HHL symptom complex to generate more specific, testable hypotheses for future research.

We focus on binaural unmasking, the auditory system’s ability to use interaural time and level differences to segregate a target sound from masking noise. This function, quantified by the Binaural Intelligibility Level Difference (BILD), depends on precise temporal processing in the auditory brainstem. It is therefore a plausible candidate for a perceptual correlate of neural desynchronization (Bharadwaj et al., 2014). While high-frequency unmasking relies primarily on interaural level differences (ILDs), which may be less affected by synchrony loss, processing broadband speech stimuli still requires robust central processing to segregate the target from the masker, a process potentially vulnerable to degraded neural input.

Our study employed a two-stage analytical approach. The first stage, a “lumping” strategy, directly tested the clinical utility of a unitary “HHL profile” by attempting to group participants based on a collection of audiometric and self-reported markers. The failure of this approach served as the motivation for our second stage, a pre-planned exploratory “splitting” strategy. This analysis was designed to investigate whether the cardinal complaints of HHL—tinnitus and SiN difficulty—are independently associated with a BILD deficit. This design allows us to move beyond asking if a deficit exists in a broadly defined “HHL group” to exploring who specifically might exhibit it, thereby generating a more refined hypothesis about the nature of these common listening complaints.

## Methods

### Participants

Thirty participants (20 Male, 10 Female; mean age = 42.7 years, SD = 14.8) were recruited. Inclusion criteria stipulated age ≥18 and proficient English. Exclusion criteria included current ear infections, significant conductive hearing loss, or known neurological conditions. Within the cohort, 17 (56.7%) reported speech perception difficulty and 13 (43.3%) reported tinnitus. All procedures were approved by the University of Southampton Institutional Research Ethics Committee (ERGO: 92186). Informed consent was obtained from all participants prior to their inclusion in the study, and their privacy rights have been observed throughout. A summary of the cohort’s demographic information is provided in Table 1.

**Table 1:** Demographic characteristics of the total participant sample.

| Characteristic | Overall Sample (N=30) |
| --- | --- |
| Age |  |
| Mean (SD) | 42.7 (14.8) years |
| Range | 23 - 71 years |
| Sex |  |
| Male | 20 (66.7%) |
| Female | 10 (33.3%) |
| Self-Reported Symptoms |  |
| Speech Difficulty | 17 (56.7%) |
| Tinnitus | 13 (43.3%) |

### Apparatus and Stimuli

Speech Reception Thresholds (SRTs) were measured using a modified British English version of the Oldenburg Matrix Test (OLMT) (Kollmeier et al., 2015) under two binaural listening conditions:

Diotic (S₀N₀): Speech and noise presented identically and in-phase to both ears. Dichotic (Antiphasic S₀Nπ): Speech in-phase, masking noise 180° out-of-phase between ears.

An auditory version of the Digit Span test was administered, and scores for the Forward and Backward sections were analyzed separately as proxies for short-term and working memory, respectively (Ryan & Lopez, 2001). We acknowledge that while complex span tasks (e.g., Reading Span) are considered the gold standard for measuring the working memory construct most relevant to speech-in-noise research (Rönnberg et al., 2013), Digit Span was used here as a more readily available proxy.

### Key Outcome Measures and Predictors

A number of derived metrics were calculated from the raw data to serve as the primary variables for analysis. The following terms and abbreviations are used consistently throughout the manuscript.

Binaural Intelligibility Level Difference (BILD): The primary measure of binaural processing, calculated as the difference in Speech Reception Thresholds (SRTs) between the diotic and dichotic listening conditions (SRT_diotic – SRT_dichotic). A larger BILD indicates more e77ective binaural unmasking.

Low-Frequency Pure-Tone Average (PTA-LF): The average hearing threshold across both ears for conventional frequencies (0.5, 1, and 2 kHz), used to characterize standard hearing sensitivity.

Extended High-Frequency Pure-Tone Average (PTA-EHF): The average hearing threshold across both ears for high and extended-high frequencies (4, 8, 11.2, and 16 kHz).

ABR Wave V/I Amplitude Ratio: A physiological measure derived from the auditory brainstem response, calculated as the amplitude of Wave V divided by the amplitude of Wave I.

Forward Digit Span and Backward Digit Span: These measures, used as proxies for short-term and working memory respectively, refer to the number of digits correctly recalled in the corresponding sub-test of the Digit Span task.

Self-Reported Speech difficulty and Self-Reported Tinnitus: These refer to the binary (Yes/No) classifications based on participant responses to direct questions about their perceptual experiences. In the context of statistical models, these are treated as categorical predictors. We acknowledge that the use of binary classifications for these complex perceptual experiences is a simplification; these data were collected as part of a broader pilot study, and future work should employ continuous, validated scales (e.g., SSQ, THI).

### Audiological and Electrophysiological Recordings

Pure-Tone Audiometry, including extended high frequencies (0.5, 1, 2, 4, 8, 11.2, and 16 kHz), was performed. Bio-LogicNavigator Pro (Natus Medical Inc., Middleton, WI, USA) was used to record auditory brainstem responses. The better ear as defined by the pure-tone audiometry average (0.5-2KHz) was the test-ear. A mastoid montage with surface electrodes (non-inverting at high forehead-Fz, inverting on ipsilateral (test ear) mastoid, ground on contralateral (non-test ear) mastoid) was used to record the evoked potential. The stimuli used was a click as evidenced to have higher sensitivity than frequency-specific stimuli (Bramhall et al., 2017) in cochlear synaptopathy and was presented at 80 dB SPL. This stimuli level is considered high enough to stimulate LSR (implicated in cochlear synaptopathy) and yet not too high to be affected by OHC dysfunction where at high stimulus levels wave I amplitude maybe affected by decreased cochlear gain (Bharadwaj et al., 2014; Bramhall et al., 2017; Gu et al., 2012). The click was 100 μs in duration and presented at a rate of 11.33 Hz via TDH-39 headphones in alternating polarity to get rid of cochlear microphonics bleeding into wave I. Participants sat in a quiet room on a reclining chair relax and were asked to close their eyes, relax and minimize movement. Electrode impedances were maintained at <5 kΩ. Two runs of 2000 artifact-free sweep responses were obtained where individual recordings exceeding 23.8 μV were rejected from the average. ABR waveforms were bandpass filtered (100–3000 Hz).

Initially, a computer program estimated wave I and V amplitude. After that, wave peaks and troughs were manually confirmed and adjusted, if necessary, by the first and second author independently. The wave amplitude was measured from the peak to the following trough. Wave V/I amplitude ratio was used as the outcome measure which helps adjust for individual differences including age, gender, hearing loss, as well as maximize the cochlear synaptopathy effect where the wave I amplitude is expected to be diminished while the wave V amplitude is to be enhanced as a result of central compensation (Prendergast et al., 2017).

### Statistical Analysis

All analyses were performed using MATLAB (R2025a).

Analysis Stage 1: “Lumping” via Grouping Approaches. This stage was designed to directly test the clinical utility of HHL as a unitary diagnostic construct. The “lumping” hypothesis posits that individuals can be meaningfully grouped into a single “HHL profile” that predicts a specific functional deficit—in this case, a reduced BILD. We tested this using two distinct methods.

Theory-Driven Classification: We first created a model based on idealized clinical profiles from the literature. Three prototype vectors were defined for Normal Hearing (NH), Mild Hearing Loss (MHL), and Hidden Hearing Loss (HHL) groups. Each participant was then assigned to the group whose prototype was “closest” in a multi-dimensional space, as determined by the minimum weighted Euclidean distance. This classification was based on six z-scored features chosen to capture the canonical definition of HHL: low- and high-frequency PTA, putative ABR markers of synaptopathy (inverted Wave I amplitude and inverted Wave V/I ratio), and the primary self-reported symptoms (’Speech Perception difficulty’ and ‘Tinnitus’). Three prototype vectors, representing the idealized z-scored profile for each group, were defined as follows: NH: [−1, −1, −1, −1, −1, −1]; MHL: [1, 1.5, 0, 0, 1, 0]; HHL: [−1, 1, 1, 1, 1.5, 1]. This approach was chosen not to create a definitive classification, but to formalize the type of clinical archetypes often used implicitly in audiological practice and test whether such a heuristic-based model has predictive utility.

Data-Driven Clustering: To address the subjectivity of the theory-driven model, we performed a confirmatory analysis using a data-driven k-means clustering algorithm. This approach makes no a priori assumptions, instead partitioning the participants into objective groups based on mathematical similarity across the same six features. The number of clusters was set to k=3 to parallel the theory-driven approach. For both the theory-driven and data-driven groupings, the same statistical test was applied: an LMM was used to assess the significance of the interaction between the derived Group and the listening Condition on SRTs. A significant interaction would indicate that at least one group exhibited a different BILD, supporting the “lumping” hypothesis.

Analysis Stage 2: Exploratory “Splitting” via Linear Mixed-Effects Modelling. In contrast to grouping, this exploratory stage sought to deconstruct the HHL complex by testing the unique contribution of individual factors. Our primary analysis employed a granular LMM to predict SRTs. The model included fixed effects for listening Condition (Diotic, Dichotic) and, crucially, the interaction of Condition with our key predictors: Self-Reported Tinnitus, Self-Reported Speech difficulty, and z-scored continuous covariates for age and PTA-EHF. To reduce the number of predictors in the model given the sample size, the z-scored Forward and Backward Digit Span scores were averaged to create a single ‘Memory’ composite variable. A random intercept for Subject accounted for the repeated-measures design. A significant interaction between a predictor and Condition indicates that the predictor specifically modulates the BILD.

Analysis Stage 3: Analysis of Cognitive and Electrophysiological Data. To directly test the relationship between the subjective speech complaint and cognitive function, independent two-sample t-tests were performed to compare the Forward Digit Span and Backward Digit Span scores between participants who did and did not report Self-Reported Speech difficulty. To investigate the relationship between clinical factors and ABR measures, we used multiple linear regression, including a targeted model to test the effect of Self-Reported Tinnitus on the ABR Wave V/I Amplitude Ratio while controlling for PTA-EHF. For all tests, an alpha level of p<.05 was considered statistically significant.

## Results

An exploratory correlation analysis (Figure 1) revealed several relationships of interest. Notably, a negative correlation was observed between BILD and Tinnitus (r = −0.40), providing an initial suggestion that participants with tinnitus tend to have a smaller binaural benefit.

**Figure 1:**
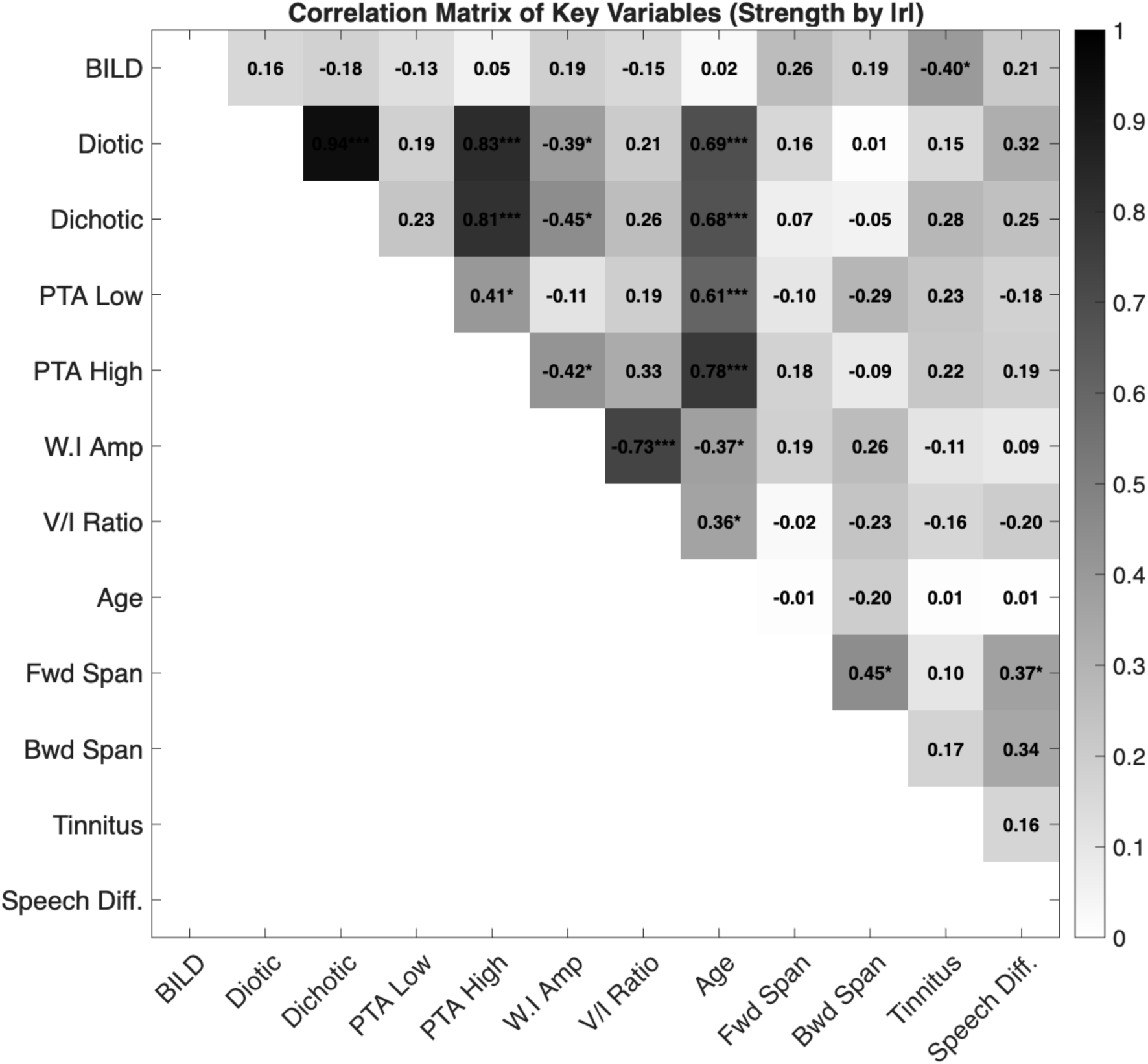
Correlation Matrix of Key Variables. Pearson correlation coefficients between the main audiometric, cognitive, and self-reported variables. The shading of each cell indicates the absolute strength of the correlation (|r|), with darker shades representing stronger correlations. The value within each cell is the correlation coefficient, r. Asterisks denote the level of statistical significance (* p < .05, ** p < .01, *** p < .001) without correction for multiple comparisons; the matrix should be interpreted as a purely exploratory guide. A notable negative correlation is observed between the presence of tinnitus and the Binaural Intelligibility Level Difference (BILD) (r = −0.40).

### Stage 1: Group Classification Analysis (“Lumping”)

Both “lumping” approaches failed to identify subgroups with a significant deficit in binaural unmasking. The theory-driven classification showed no significant Group x Condition interaction (F(2,30)=0.23, p= 0.793), a result visualized in Figure 4 (left panel). This null result was replicated using the data-driven k-means clustering approach (F(2,30)=0.61, p= 0.549), which also failed to produce well-separated. A principal component analysis of the features used for classification further highlighted the poor separation between groups (Figure 3). The first two principal components accounted for only 36.2% and 23.7% of the variance, respectively. This low cumulative variance demonstrates that these common clinical markers do not combine to produce well-defined patient archetypes. Collectively, these results indicate that in our sample, a “lumped” HHL profile was not a useful predictor of a functional BILD deficit.

**Figure 2:**
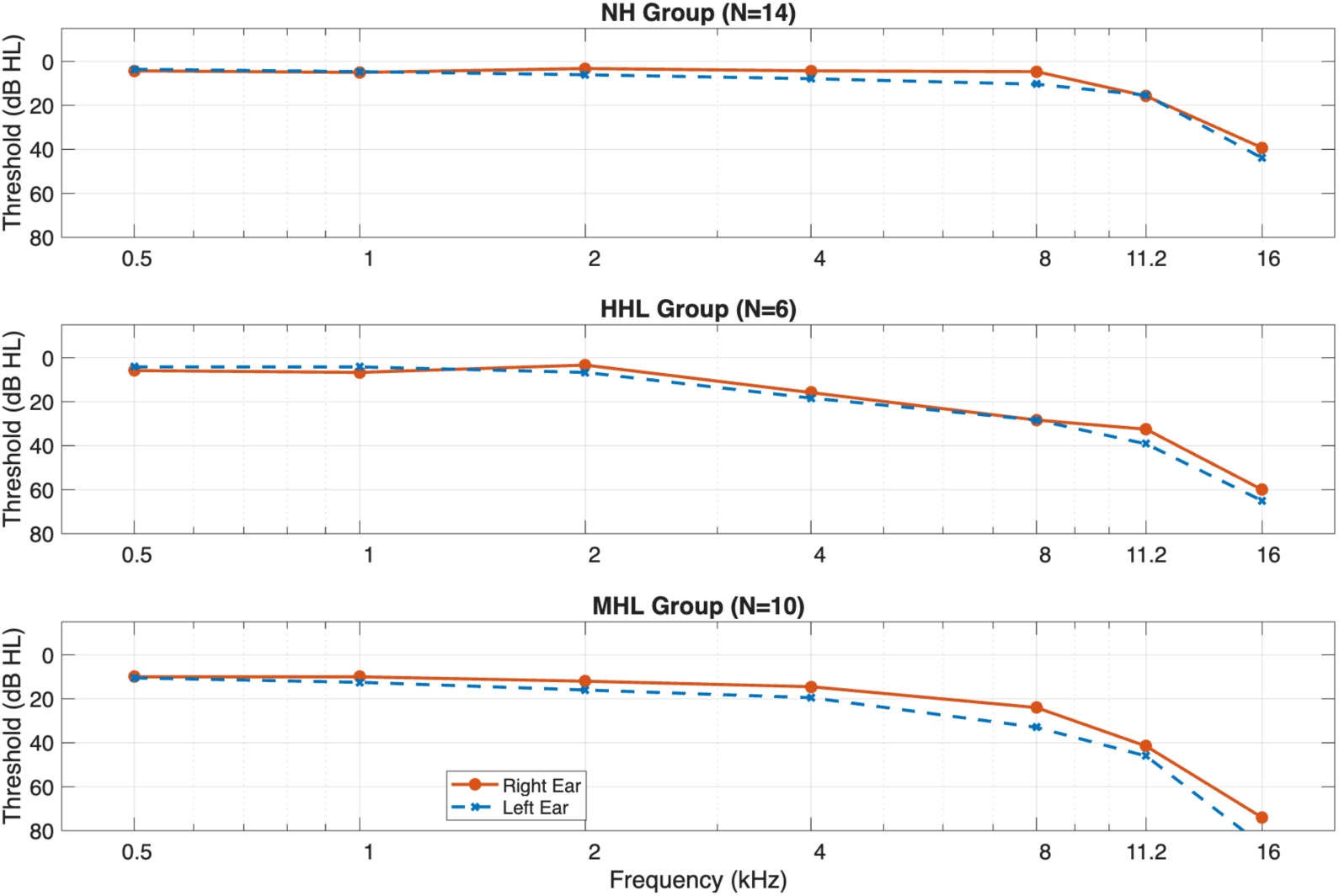
Mean audiograms for participant groups based on theory-driven classification. The plots show the mean pure-tone thresholds (in dB HL) across extended high frequencies for the right (solid red line) and left (dashed blue line) ears. Participants were classified into Normal Hearing (NH, N=14), Hidden Hearing Loss (HHL, N=6), and Mild Hearing Loss (MHL, N=10) groups based on their distance from idealized prototypes.

**Figure 3:**
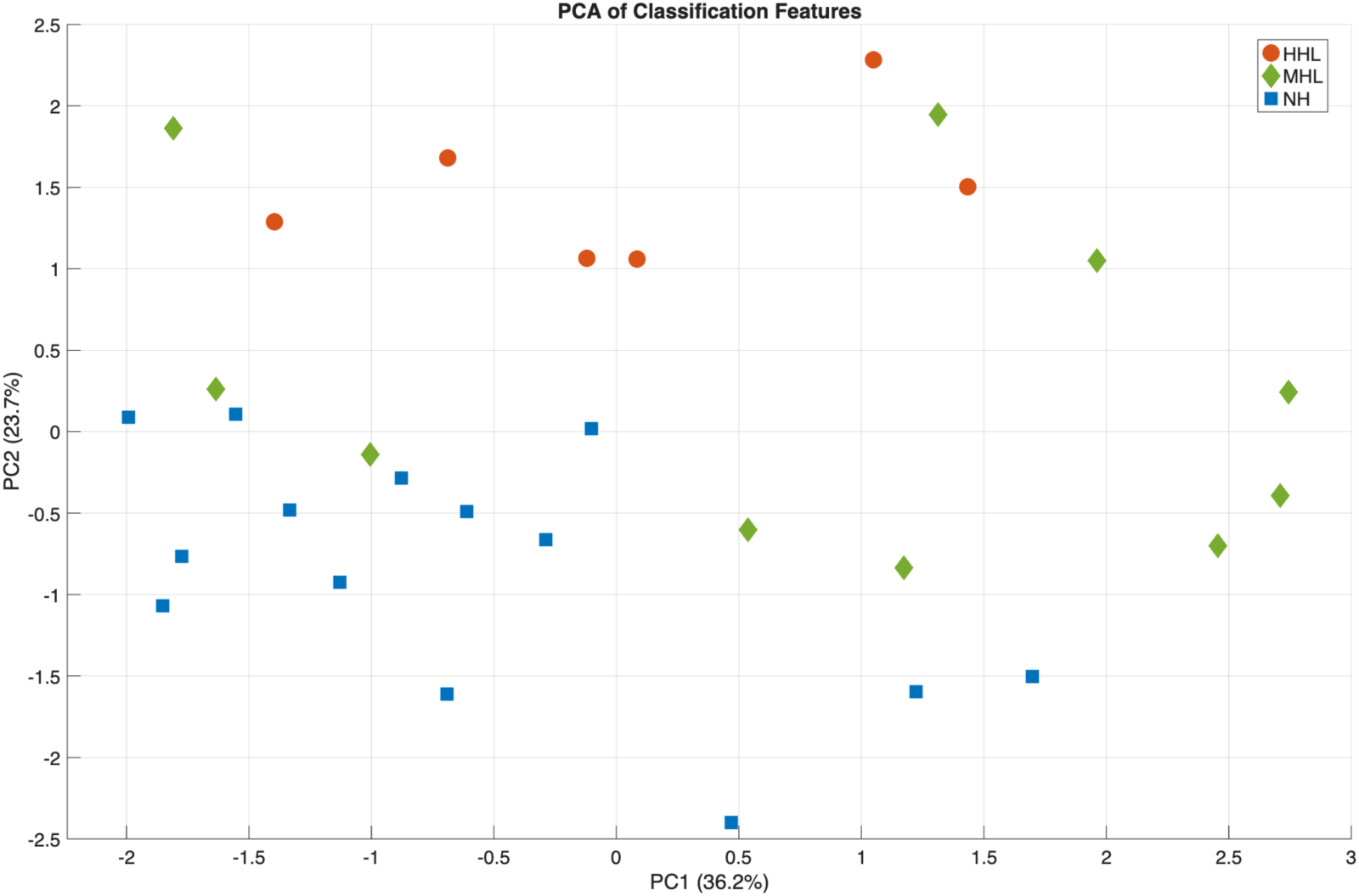
Principal Component Analysis (PCA) of classification features. A scatter plot showing the distribution of participants along the first two principal components derived from the six features used for classification. Each point represents an individual participant, with the symbol and colour indicating their assigned group from the theory-driven classification (Squares: NH, Diamonds: MHL, Circles: HHL). The percentage of total variance explained by each component is shown on the axis labels (PC1: 36.2%, PC2: 23.7%).

**Figure 4:**
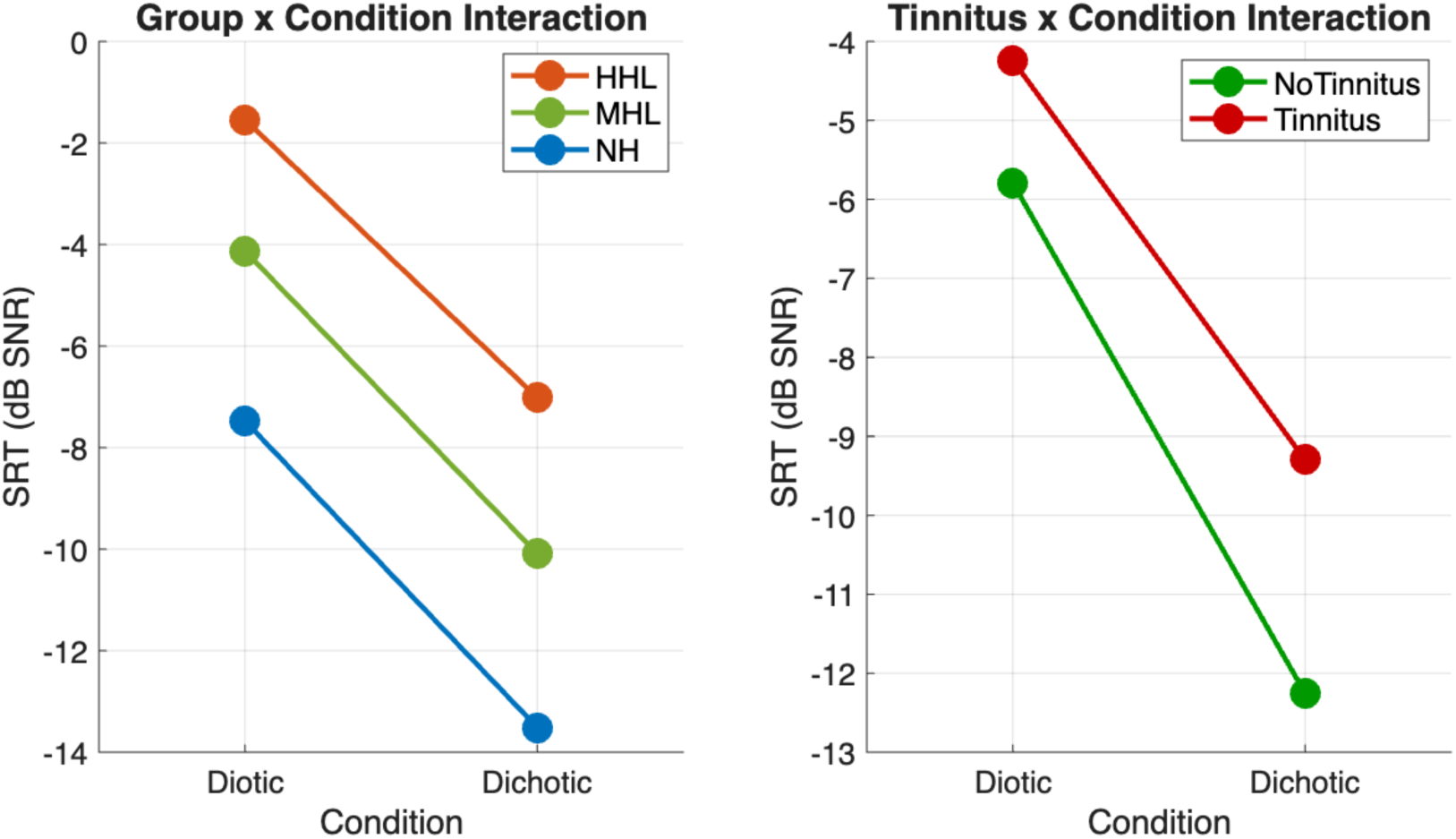
Interaction plots showing the effect of grouping and tinnitus on binaural unmasking. The plots show the mean Speech Reception Thresholds (SRTs) under diotic (S₀N₀) and dichotic (S₀Nπ) listening conditions. The slope of the line represents the Binaural Intelligibility Level Difference (BILD). (Left Panel) The non-significant interaction between the theory-driven groups (NH, HHL, MHL) and listening condition (p=0.793), indicating that the BILD did not differ significantly between these groups. (Right Panel) The significant interaction between tinnitus and listening condition (p=0.009), demonstrating that participants with tinnitus (red line) had significantly poorer SRTs in the dichotic condition, resulting in a smaller BILD compared to those without tinnitus (green line).

### Stage 2: Exploratory Linear Mixed-Model Analysis (“Splitting”)

The failure of the grouping analysis motivated the exploratory LMM to test for contributions from individual factors (Table 2). The central finding of this exploratory analysis was a significant interaction between Tinnitus and Condition (β = 1.57, p = 0.009, 95% CI [0.42, 2.72]), indicating that participants with tinnitus exhibited a significantly smaller BILD. This interaction is illustrated in Figure 4 (right panel), which clearly shows a reduced benefit from dichotic listening in the tinnitus group. The interaction between Self-Reported Speech difficulty and Condition did not reach statistical significance (p=0.086). Given the study’s limited power, this may represent a Type II error; however, in the context of our exploratory framework, it provides one half of an emergent dissociation. Figure 5 visually confirms that only the Tinnitus:Condition interaction’s confidence interval does not overlap with zero.

**Figure 5:**
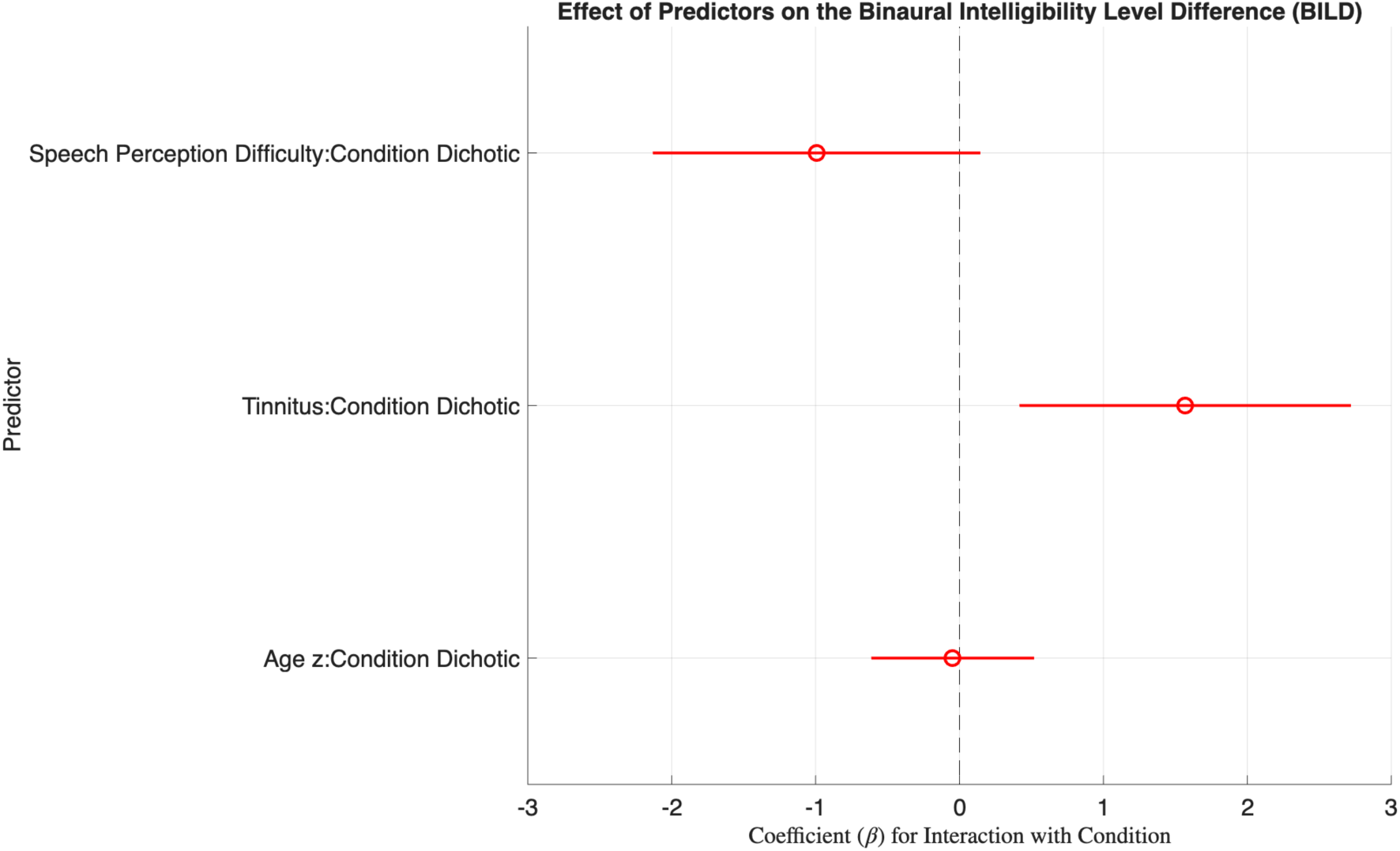
Coefficient plot for key interaction terms from the Linear Mixed-Effects Model (LMM). The plot displays the estimated coefficients (β) and their corresponding 95% confidence intervals for the interaction between listening condition (Dichotic) and three predictors: self-reported speech perception difficulty, tinnitus, and age. A predictor is considered to have a significant effect on the BILD if its confidence interval does not overlap with zero (dashed vertical line). Only the Tinnitus:Condition interaction is statistically significant, with its coefficient of 1.57 dB indicating a reduction in binaural unmasking for participants with tinnitus.

**Table 2:** LMM Predicting SRTs (dB SNR) from Specific Clinical and Audiometric Factors.

| Fixed Effect | Est. | SE | tStat | p | Lower CI | Upper CI |
| --- | --- | --- | --- | --- | --- | --- |
| Intercept | -6.36 | 0.92 | -6.94 | <.001 | -8.20 | -4.52 |
| Speech Diff. | 2.25 | 1.14 | 1.97 | 0.054 | -0.05 | 4.55 |
| Tinnitus | -0.23 | 1.10 | -0.21 | 0.083 | -2.44 | 1.97 |
| Age (z) | 1.00 | 0.85 | 1.18 | 0.244 | -0.71 | 2.72 |
| PTA EHF (z) | 3.27 | 0.87 | 3.78 | <.001 | 1.53 | 5.01 |
| Memory (z) | -0.19 | 0.56 | -0.35 | 0.731 | -1.31 | 0.92 |
| Condition (Dichotic) | -5.96 | 0.46 | -13.07 | <.001 | -6.88 | -5.05 |
| Speech Diff.:Dichotic | -0.99 | 0.57 | -1.75 | 0.086 | -2.13 | 0.14 |
| Tinnitus:Dichotic | 1.57 | 0.57 | 2.74 | 0.009 | 0.42 | 2.72 |
| Age (z):Dichotic | -0.05 | 0.28 | -0.17 | 0.867 | -0.61 | 0.52 |

### Stage 3: Analysis of Cognitive and Electrophysiological Data

When cognitive performance was compared, participants reporting Self-Reported Speech difficulty (N=17) had significantly lower scores on the forward digit span test (a proxy for short-term memory) than those who did not (N=13) (t(28) = −2.09, p = 0.046). A similar, non-significant, trend was observed for the backward digit span test (a proxy for working memory) (t(28) = −1.94, p = 0.063). This finding provides the second half of the dissociation suggested by the LMM analysis.

In the ABR analysis, the full regression model showed no significant predictors of the Wave V/I ratio. In our targeted follow-up analysis to test the effect of tinnitus while controlling for hearing loss, PTA-EHF was a significant predictor of the Wave V/I ratio (p= 0.041), but Tinnitus was not (p= 0.177).

## Discussion

The primary contribution of this work is not a definitive conclusion, but the articulation of a motivated, testable hypothesis derived from an explicit analytical strategy. We first demonstrated that “lumping” participants into a unitary HHL profile, using either theory-driven or data-driven methods, failed to predict a functional deficit in binaural unmasking. This null result is a key finding, as it challenges the utility of this broad diagnostic label and provided the direct rationale for our subsequent exploratory “splitting” analysis. This second analysis generated a clear hypothesis of a sensory-cognitive dissociation: in our sample, tinnitus was specifically associated with a reduced BILD, whereas the complaint of speech-in-noise difficulty was not, but was instead associated with poorer short-term memory.

### Interpreting the Findings: A Tentative Sensory-Cognitive Hypothesis

We propose a sensory-cognitive framework to interpret this pattern, stressing that it is a hypothesis generated from an underpowered, exploratory analysis.

The tinnitus-associated BILD deficit appears linked to a bottom-up, sensory-level processing issue. One potential mechanism is a form of binaural interference. If tinnitus arises from aberrant central gain following peripheral deafferentation (Schaette & McAlpine, 2011), this chronic neural hyperactivity may act as disruptive ‘internal noise’, contaminating the precise temporal computations in the brainstem required for unmasking. An equally plausible alternative is that the deficit is cognitive. The reduced BILD may not reflect a degraded sensory signal, but rather a diminished ability to allocate the top-down attentional resources required by the more demanding dichotic condition—a process known to be taxed by the constant presence of tinnitus (Husain & Schmidt, 2014)

In contrast, the complaint of general SiN difficulty appears, in our data, to be linked to top-down cognitive factors. The association we found between this complaint and short-term memory aligns with the “Framework for Understanding Effortful Listening” (FUEL), which posits that hearing difficulty can arise from the increased “cognitive energy” required to parse a degraded or complex auditory scene (Pichora-Fuller et al., 2016). For these individuals, their complaint may not reflect a specific sensory deficit like poor binaural processing, but rather a limitation in the general cognitive resources available for effortful listening.

Crucially, the tinnitus-related BILD deficit was not reflected in our click-evoked ABR measures. This aligns with a growing body of literature demonstrating the insensitivity of the click-ABR for detecting subtle neural deficits (Bramhall et al., 2017; Marmel et al., 2020) and underscores the need for more advanced functional measures to probe the neural correlates of these distinct perceptual complaints.

### Limitations and Future Directions

This study has significant limitations. The primary limitation is the small sample size (N=30), which provides limited statistical power and means our findings must be considered preliminary. The exploratory “splitting” analysis that generated our main hypothesis is, by its nature, post-hoc and carries an inflated risk of HARKing.

Furthermore, our methodology has weaknesses. The operationalization of tinnitus and speech difficulty relied on binary questions, which is a gross simplification. Our cognitive measure (Digit Span) was a proxy, not a gold-standard complex span task. The lack of noise-exposure history for participants prevents any firm conclusions about cochlear synaptopathy, and the manual ABR analysis was not performed blind.

These limitations define a clear roadmap for future research. The sensory-cognitive dissociation proposed here must be tested in a large-scale, pre-registered replication study. Such a study should use continuous, validated questionnaires (e.g., THI, SSQ), a comprehensive cognitive battery including complex span tasks, and more sensitive electrophysiological measures (e.g., the speech-evoked ABR or envelope following responses) to establish the convergent and discriminant validity of these distinct profiles.

## Conclusion

The results of this exploratory study challenge the clinical utility of a unitary HHL profile for predicting deficits in binaural unmasking. By contrasting broad grouping methods with a more granular exploratory model, we generated a specific, testable hypothesis of a sensory-cognitive dissociation within the HHL symptom complex. Our preliminary data suggest that tinnitus may be linked to a functional deficit in binaural sensory processing, whereas the complaint of speech difficulty may be associated with cognitive factors. This framework, however, is derived from post-hoc analysis of an underpowered study and must be treated with appropriate caution. Its value lies in providing a motivated hypothesis that now requires rigorous investigation.

## Data Availability

All data produced in the present study are available upon reasonable request to the authors

## Acknowledgements

We thank the RNID (Royal National Institute for Deaf People) in the UK for funding. We also thank our students Eloisa Kattenhorn and Dewan Chowdhury for help with data acquisition.

## Notes

### Competing Interest Statement

The authors have declared no competing interest.

### Author Declarations

All procedures were approved by the University of Southampton Institutional Research Ethics Committee (ERGO: 92186).

## References

Bharadwaj, H. M., Verhulst, S., Shaheen, L., Charles Liberman, M., & Shinn-Cunningham, B. G. (2014). Cochlear neuropathy and the coding of supra-threshold sound. Frontiers in Systems Neuroscience, 8(FEB). 10.3389/fnsys.2014.00026

Bramhall, N. F., Konrad-Martin, D., McMillan, G. P., & Griest, S. E. (2017). Auditory Brainstem Response Altered in Humans With Noise Exposure Despite Normal Outer Hair Cell Function. Ear and Hearing, 38(1), e1–e12. 10.1097/AUD.0000000000000370

Gu, J. W., Herrmann, B. S., Levine, R. A., & Melcher, J. R. (2012). Brainstem auditory evoked potentials suggest a role for the ventral cochlear nucleus in tinnitus. Journal of the Association for Research in Otolaryngology : JARO, 13(6), 819–833. 10.1007/S10162-012-0344-1

Husain, F. T., & Schmidt, S. A. (2014). Using resting state functional connectivity to unravel networks of tinnitus. In Hearing Research (Vol. 307, pp. 153–162). 10.1016/j.heares.2013.07.010

Kollmeier, B., Warzybok, A., Hochmuth, S., Zokoll, M. A., Uslar, V., Brand, T., & Wagener, K. C. (2015). The multilingual matrix test: Principles, applications, and comparison across languages: A review. International Journal of Audiology, 54 Suppl 2, 3–16. 10.3109/14992027.2015.1020971

Liberman, M. C., & Kujawa, S. G. (2017). Cochlear synaptopathy in acquired sensorineural hearing loss: Manifestations and mechanisms. Hearing Research, 349, 138–147. 10.1016/J.HEARES.2017.01.003

Marmel, F., Cortese, D., & Kluk, K. (2020). The ongoing search for cochlear synaptopathy in humans: Masked thresholds for brief tones in Threshold Equalizing Noise. Hearing Research, 392. 10.1016/j.heares.2020.107960

Moser, T., & Starr, A. (2016). Auditory neuropathy — neural and synaptic mechanisms. Nature Reviews Neurology 2016 12:3, 12(3), 135–149. 10.1038/nrneurol.2016.10

Pichora-Fuller, M. K., Kramer, S. E., Eckert, M. A., Edwards, B., Hornsby, B. W. Y., Humes, L. E., Lemke, U., Lunner, T., Matthen, M., Mackersie, C. L., Naylor, G., Phillips, N. A., Richter, M., Rudner, M., Sommers, M. S., Tremblay, K. L., & Wingfield, A. (2016). Hearing Impairment and Cognitive Energy: The Framework for Understanding Effortful Listening (FUEL). Ear and Hearing, 37 Suppl 1, 5S–27S. 10.1097/AUD.0000000000000312

Prendergast, G., Millman, R. E., Guest, H., Munro, K. J., Kluk, K., Dewey, R. S., Hall, D. A., Heinz, M. G., & Plack, C. J. (2017). Effects of noise exposure on young adults with normal audiograms II: Behavioral measures. Hearing Research, 356, 74–86. 10.1016/J.HEARES.2017.10.007

Rönnberg, J., Lunner, T., Zekveld, A., Sörqvist, P., Danielsson, H., Lyxell, B., Dahlström, Ö., Signoret, C., Stenfelt, S., Pichora-Fuller, M. K., & Rudner, M. (2013). The Ease of Language Understanding (ELU) model: Theory, data, and clinical implications. Frontiers in Systems Neuroscience, 7(JUNE), 48891. 10.3389/FNSYS.2013.00031/ENDNOTE

Ryan, J. J., & Lopez, S. J. (2001). Wechsler Adult Intelligence Scale-III. Understanding Psychological Assessment, 19–42. 10.1007/978-1-4615-1185-4_2

Schaette, R., & McAlpine, D. (2011). Tinnitus with a normal audiogram: physiological evidence for hidden hearing loss and computational model. The Journal of Neuroscience : The Official Journal of the Society for Neuroscience, 31(38), 13452–13457. 10.1523/JNEUROSCI.2156-11.2011

